# Ultrasensitive detection of *Bacillus anthracis* by real time PCR targeting a polymorphism in multi-copy 16S rRNA genes and their transcripts

**DOI:** 10.1101/2021.09.20.21263746

**Authors:** Peter Braun, Martin Duy-Thanh Nguyen, Mathias C. Walter, Gregor Grass

## Abstract

The anthrax pathogen *Bacillus anthracis* poses a significant threat to human health. Identification of *B. anthracis* is challenging because of the bacterium’s close genetic relationship to other *Bacillus cereus* group species. Thus, molecular detection is founded on species-specific PCR targeting single-copy genes. Here, we validated a previously recognized multi-copy target, a species-specific SNP present in 2-5 copies in every *B. anthracis* genome analyzed. For this, a hydrolysis probe-based real time PCR assay was developed and rigorously tested. The assay was specific as only *B. anthracis* DNA yielded positive results, was linear over 9 log_10_ units and was sensitive with a limit of detection (LoD) of 2.9 copies/reaction. Though not exhibiting a lower LoD than established single copy PCR targets (*dhp61* or *PL3*), the higher copy number of the *B. anthracis*–specific 16S rRNA gene allele afforded ≤2 unit lower threshold (Ct) values. To push the detection limit even further, the assay was adapted for reverse transcription PCR on 16S rRNA transcripts. This RT-PCR assay was also linear over 9 log_10_ units and was sensitive with a LoD of 6.3 copies/reaction.In a dilution-series of experiments, the 16S RT-PCR assay achieved a thousand-fold higher sensitivity than the DNA-targeting assays. For molecular diagnostics, we recommend a real time RT-PCR assay variant in which both DNA and RNA serve as templates (thus, no requirement for DNase treatment). This will at least provide results equaling the DNA-based implementation if no RNA is present but will be superior even at the lowest residual rRNA concentrations.

## 1. Introduction

Within the genus *Bacillus* the notorious anthrax pathogen *Bacillus anthracis* poses the greatest risk for humans, mammal livestock and wildlife. [1]. Other *Bacillus* spp. such *as B. cereus* or *B. thuringiensis* may also have pathogenic traits related to food-poisoning, infections in immunocompromised persons or production of insecticides [2]. Yet only *B. anthracis* (and a few *B. anthracis*-like bacilli) features a unique suite of pathogenicity factors rendering the endospore-forming bacterium a first-rate biothreat agent. These factors are encoded on two plasmids called pXO1 and pXO2, respectively. Plasmid pXO1 encodes the anthrax toxin genes producing the lethal toxin (gene products of *pagA* and *lef*) and edema toxin (gene products of *pagA* and *cya*) [1]. These toxins damage host cells on various levels [3]. Plasmid pXO2 harbors the capsule genes endowing the pathogen with a poly-glutamyl capsule which helps evading host immune response [1, 4]. Phylogenetically, *B. anthracis* belongs to the very closely related *Bacillus cereus sensu lato* group. Besides the better-known species *B. cereus sensu stricto, B. anthracis* or *B. thuringiensis*, the group also comprises several other familiar species such as *B. weihenstephanensis, B. mycoides, B. cytotoxicus* and a variety of lesser-characterized members [5].

In the past, the high degree of genetic relatedness to several *B. cereus s. l*. strains has rendered molecular diagnostics of *B. anthracis* challenging (e.g., by polymerase chain reaction assays, PCR). One would think it should be straight forward to identify *B. anthracis* by detecting genetic marker genes (typically *pagA, lef, cya, capB* or *capC*) [6-8] on one or both of its virulence plasmids. Identifying these genes, however, only verifies the presence of these plasmids. This is relevant because several *B. cereus s. l*. isolates are documented to possess very similar virulence plasmids but not necessarily all of these belong to the species *B. anthracis*. Further, there are *B. anthracis* strains that lack one or both virulence plasmids. Species-specific molecular identification of *B. anthracis* is achieved by targeting a small number of validated chromosomal targets. These targets comprise sections of genes such as *dhp61* (*BA_5345*; [9]), *PL3* (*BA_5358*; [6]) or mutations characterized as single nucleotide polymorphisms (SNPs) e.g., in the *rpoB* [7] or the *plcR* [10] gene, respectively. A comprehensive overview of suitable and less ideal specific markers for *B. anthracis* has been provided previously [11]. Notwithstanding, the advantage of assaying for pXO1 or pXO2 markers over chromosomal ones is that the plasmid markers occur as multi-copy genes (since the virulence plasmids are present in more than one copy per cell) [12]. Large-scale genomic sequencing revealed that in *B. anthracis* plasmids pXO1 and pXO2 (with their respective PCR-marker genes) are present on average in 3.86 and 2.29 copies, respectively [13]. Conversely, no multi-copy chromosomal marker has been employed for *B. anthracis* detection thus far.

Likewise, ribosomal RNA (particularly 16S rRNA) has not yet been routinely used for identification and detection of *B. anthracis* even though rRNA molecules are generally the most abundant ribonucleic acid entities in cells constituting up to approximately 80% of total RNA [14]. In fact, copies of 16S rRNA transcripts per cell as constituents of ribosomes, number in many thousands e.g., in *E. coli* the number of ribosomes per cell ranges from 8 × 10^3^ at a doubling time of 100 min to 7.3 × 10^4^ at a doubling time of 20 min [15]. Even in stationary culture, a single *E. coli* bacterium contains about 6.5 × 10^3^ copies of ribosomes [16]. Phylogenetically closer to *B. anthracis* than *E. coli* is *Bacillus licheniformis*. For this bacillus the average number of ribosomes per cell was calculated at 1.25 × 10^4^, 3.44 × 10^4^, or 9.2 × 10^4^ in cultures growing at 37°C with generation times of 120, 60 and 35 min, respectively [17]. While these numbers are well in agreement, somewhat lower numbers of 9 × 10^3^ ribosomes have been determined for exponentially growing cells of *Bacillus subtilis* [18]. While unexplored for *B. anthracis*, bacterial detection using rRNA genes and transcripts has been successfully harnessed to challenge previous limits of detection (LoD) for other pathogens [19-21].

In this study, we introduce a species-specific multi-copy chromosomal PCR marker of *B. anthracis*. This marker is represented by a unique SNP within a variable number of loci of the multi-copy 16S rRNA gene in this organism. Though the 16S rRNA gene sequences feature a very high degree of identity among the *B. cereus s*.*l*. group species [22], this SNP has previously been identified as unique and present in all publicly available *B. anthracis* genomic data [23-25]]. Since all 16S rRNA gene copies harboring the SNP have 100% sequence identity, this specific sequence variation represents a distinct 16S rRNA gene allele named 16S-BA-allele. For simplification, all other 16S rRNA gene alleles lacking the sequence variation were named 16S-BC-allele. The relative abundance of these 16S-BA- and –BC-alleles were recently quantified in 959 *B. anthracis* isolates [25]. Here we also harnessed this SNP to develop a *B. anthracis* specific reverse transcription (RT) real time PCR assay. This approach brings the multi-copy marker concept for *B. anthracis* up to a new level owed to the excess numbers of ribosomes (and thus 16S rRNA moieties) in relation to chromosomes within a *B. anthracis* cell.

## 2. Materials and Methods

### 2.1. Bacterial culture, inactivation and DNA samples for quality assessment

*B. anthracis* strains and other Bacilli were cultivated at 37°C on tryptic soy agar plates (TSA, Merck KGaA, Darmstadt, Germany). Bacteria comprising the negative panel (Supplementary Table S1) were grown on appropriate agar media (with 10% CO_2_ atmosphere where required) at 37°C until colonies emerged. Risk group 3 (RG-3) *B. anthracis* strains were cultivated in the biosafety level 3 (BSL-3) facilities at the Bundeswehr Institute of Microbiology (IMB) and then chemically inactivated before further use [26]. RG-2 strains of endospore formers were inactivated by resuspending a loop of colony material in aqueous peracetic acid solution (4% Terralin PAA, Schülke & Mayr GmbH, Norderstedt, Germany) [26]. All other bacterial cultures were inactivated by 70% (v/v) ethanol. Ring trial *B. anthracis* DNA samples published in [27] were obtained from Instant (Düsseldorf, Germany).

### 2.2. Isolation of DNA, RNA and nucleic acid quantification

Bacterial DNA and RNA was isolated using MasterPure™ Gram Positive DNA Purification kit (Lucigen, Middleton, WI, USA). For RNA (+DNA) isolation, RNase treatment was omitted. DNA and RNA concentrations were quantified using the Qubit dsDNA HS Assay or RNA HS Assay kits (ThermoFisher Scientific, Darmstadt, Germany) according to the manufacturers’ protocols. DNA and RNA (+DNA) preparations were stored at -20°C and -80°C, respectively, until further use.

### 2.3. Design and in silico bioinformatic analysis of primer and probe DNA sequences

All relevant DNA sequence data for oligonucleotide design were retrieved from public databases (NCBI). Primer and probe DNA oligonucleotides [25] were designed with Geneious Prime (Biomatters, USA). *In silico* specificity analysis was performed by probing each primer and probe nucleotide sequences against the NCBI nt databases using BLASTN for short input sequences (Primer BLAST) [28]. The two amplification oligonucleotide primers target a consensus region within the 16S rRNA genes on the chromosome of *B. cereus s*.*l*. species (Table 1) including *B. anthracis*. The two oligonucleotide probes (Table 1) feature the centrally located discriminatory SNP (pos. 1110 in *B. anthracis* strain Ames Ancestor, NC_007530) [23, 24]. These probes thus either match the allele unique for *B. anthracis* (named 16S-BA-allele; with an adenine, A at the SNP position) or the general 16S-BC-allele (guanine, G at the SNP position), respectively (the two alleles are depicted in Supplementary Figure S1). Due to placement and length restrictions related to another non-discriminatory SNP (pos. 1119) each probe was amended with locked nucleic acids (LNA). LNA are modified nucleic acids in which the sugar is conformationally locked. This rigidity causes exceptional hybridization affinity through stable duplexes with DNA and RNA [29] eventually improving mismatch discrimination in SNP genotyping studies. LNA probes as well as primers were purchased from TIB MolBiol (Berlin, Germany).

**Table 1:**
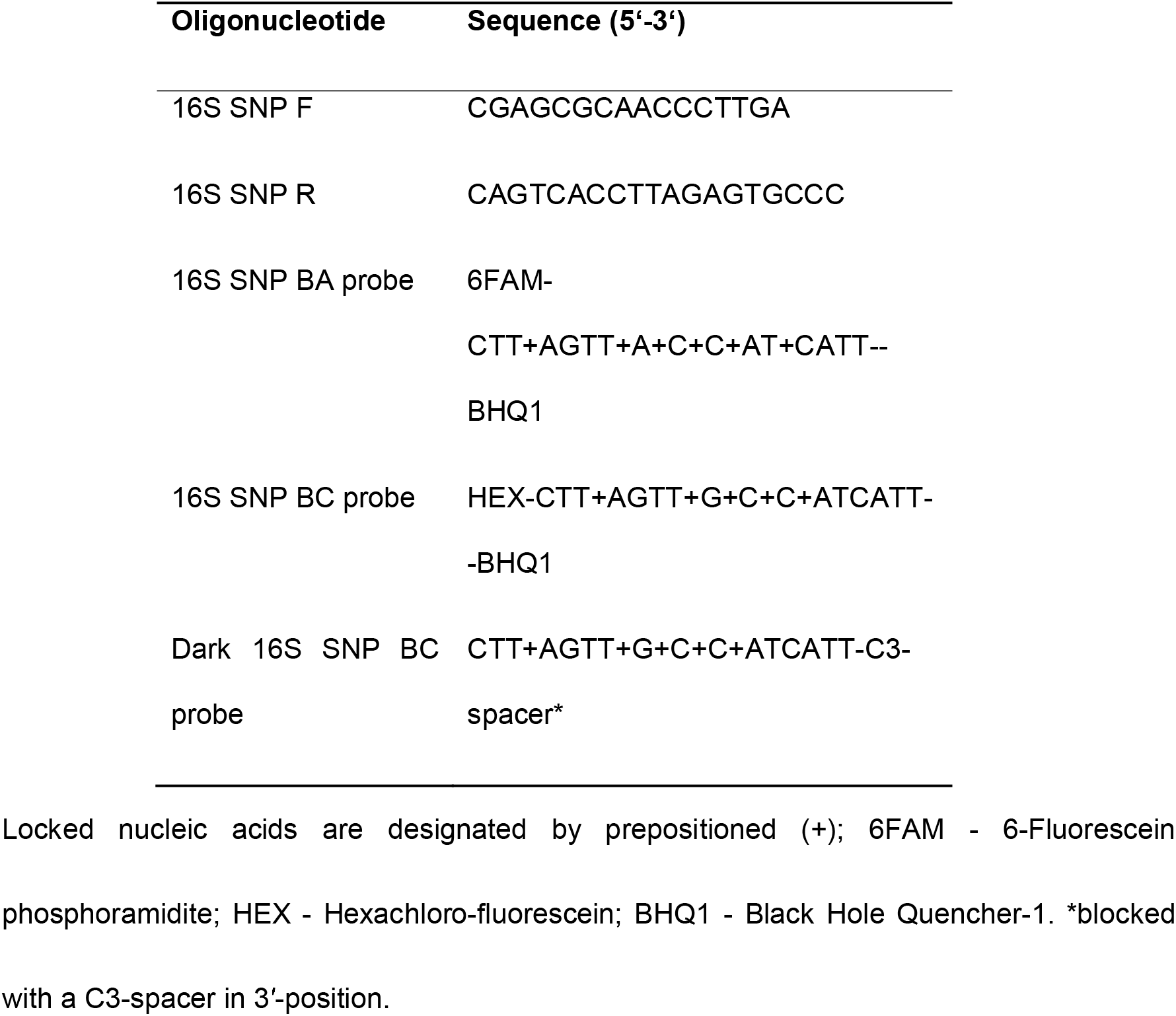
primers and probes.

### 2.4. Real-time and reverse transcription PCR conditions

All (pseudo) duplex real-time PCR amplifications were performed in reaction mixtures of a final volume of 20 µl containing 2 μL LightCycler® FastStart DNA Master HybProbe mix (Roche Diagnostics, Mannheim, Germany), 5 mM MgCl_2_, 0.5 μM of each primer, 0.25 µM of 16S SNP BA probe, 0.75 µM of (dark) 16S SNP BC probe, and various quantities of template DNA template. All reactions were performed on a LightCycler 480 real-time PCR system fitted with color compensation (Roche Diagnostics, Mannheim, Germany). The optimized amplification conditions were: 95°C for 10 min, and then 45 consecutive cycles of first 15 s at 95°C and then 20 s at 62°C, followed by 20 s at 72°C.

Reverse transcription PCR reaction mixtures contained 7.4 µl LightCycler® 480 RNA Master Hydrolysis Probes mix, 1.3 µl Activator, 1 µl Enhancer (Roche Diagnostics, Mannheim, Germany), 0.5 μM of each primer, 0.25 µM of 16S SNP BA probe, 0.75 µM of (Dark) 16S SNP BC probe, a variable volume of RNA and/or DNA template. Finally nuclease-free water (Qiagen, Hilden, Germany) was added to a final volume of 20 µl. Using the LightCycler 480 real-time PCR system (Roche Diagnostics, Mannheim, Germany) reverse transcription was performed at 63°C for 3 min followed by an activation step at 95 °C for 30 s and 45 cycles of 95 °C for 15 s, 62 °C for 20 s and 72 °C for 1 s.

A fluorescent signal 10-fold higher than the standard deviation of the mean baseline emission was counted a positive detection. Samples were tested in triplicate (unless noted otherwise) and data recorded as Cycle thresholds (Ct) with Ct defined as the PCR cycle at which the fluorescent intensity raised above the threshold [30].

### 2.5. Droplet digital PCR (ddPCR) and reverse transcription (RT) ddPCR

All DNA and RNA templates used for real-time and reverse transcription PCR were quantified by ddPCR and RT ddPCR, respectively. A 20 μl ddPCR reaction mixture consisted of 10 μl ddPCR Supermix for Probes (Bio-Rad Laboratories, Munich, Germany), 0.9 µM of each primer, 0.15 µM of each probe and 5 µl of template DNA. RT-ddPCR reaction mixtures comprised of 5 μl One-Step RT-ddPCR Advanced Supermix for Probes (Bio-Rad, Munich, Germany), 2 µl of Reverse Transcriptase (Bio-Rad, Munich, Germany; final concentration 20 U/µl), 0.6 µl of DTT (Bio-Rad, Munich, Germany; final concentration 10 nM),), 0.9 µM of each primer, 0.15 µM of each probe and 5 µl of template RNA. Droplets were generated using a QX200 ddPCR droplet generator (Bio-Rad, Munich, Germany). PCR amplification for both assays was performed on the Mastercycler Gradient (Eppendorf, Hamburg, Germany) with the following conditions:

Initial reverse transcription was carried at 48°C for 60 min (only for RT-ddPCR). Enzyme activation at 95°C for 10 min was followed by 40 cycles of a denaturation at 94°C for 30 s and annealing/extension at 58°C for 1 min. Before the samples were cooled to 4°C a final enzyme inactivation was carried out at 98°C for 10 min. Cooling and heating ramp rate was set to 2°C/s for all steps. After PCR runs, droplets were analyzed using the QX100 Droplet Reader (Bio-Rad, Munich, Germany) and absolute target concentrations of each sample were calculated using Quantasoft Pro Software (Bio-Rad, Munich, Germany).

### 2.6. Generation of PCR positive controls from reference plasmids harboring 16S-BA- or BC-allele fragments

Though we generally used genomic DNA from *B. cereus* or *B. anthracis*, respectively, for PCR testing and validation, generic positive control reference plasmids for either allele, the *B. anthracis*-specific 16S-BA-allele or the *B. cereus*-specific 16S-BC-allele were constructed. For this, a PCR-amplicon was generated from *B. anthracis* Ames DNA with primers 16S SNP F and 16S SNP R using Platinum™ Taq DNA Polymerase High Fidelity (ThermoFisher Scientific, Darmstadt, Germany). This DNA comprises a mixture of both alleles in a ratio of 4 to 7 [25]. The PCR-amplicon was analyzed on agarose gel electrophoresis, a band of the expected size (57 bp) cut from the gel and gel-purified using QIAquick Gel Extraction kit (QIAGEN, Hilden Germany). PCR products were ligated into pCR2.1 TOPO vector (ThermoFisher Scientific, Darmstadt, Germany) using TOPO TA Cloning kit (Thermo Scientific, Darmstadt, Germany) and transformed into One Shot TOP10 chemically competent cells (ThermoFisher Scientific, Darmstadt, Germany) according to the manufacturer’s protocol. Several recombinant plasmids isolated from different clones were sequenced (Eurofins Genomics Germany, Ebersberg, Germany) in order to obtain plasmids harboring either the 16S-BA-allele or the 16S-BC-allele. From these plasmids, PCR products were generated using primers M13 F and M13 R, which contained the target region for the 16S rRNA SNP-PCR with either the 16S-BA- or BC-allele. After purification with QIAquick PCR purification kit (QIAGEN, Hilden Germany) PCR products were quantified using digital PCR and diluted as required.

### 2.7. Determination of the specificity (inclusivity/exclusivity) of the B. anthracis 16S rRNA allele assay

PCR specificity for the 16S rRNA SNP assay was assessed by verifying amplification of DNA containing or lacking respective markers. “Inclusivity” was evaluated by (exponential) amplification above threshold levels obtained with template DNA comprising the markers’ sequences. Vice versa, “exclusivity” was confirmed by lack of amplification of genomic DNA from *B. cereus s*.*l*. strains reported to lack the particular 16S-BA-allele, but also may harbour the alternative 16S-BC-allele or include no-template negative controls (NTC). Positive PCR results were further analysed via agarose gel electrophoresis demonstrating a single band with a molecular weight corresponding to the predicted size of the 16S rRNA SNP-PCR amplicon (note: this cannot differentiate between the two alternative SNP states in the 16S rRNA gene alleles).

### 2.8. Dynamic linear range, PCR efficiency and limit of detection

The dynamic linearity of the PCR assays were determined over a 9 log_10_ concentration range for DNA (real-time PCR) and RNA (RT-PCR) templates. Each dilution was assayed 6-fold, and analysis for linearity and PCR-efficiency (E) was performed from the plot of the Ct’s versus the logarithm of the target concentrations [31]. The sensitivity of the PCR assay was expressed as the limit of detection (LoD) of 16S rRNA SNP genome or transcript copies. LoD was formally defined as the concentration permitting detection of the analyte at least 95% of the time. For this, DNA fragments comprising the 16S rRNA SNP were diluted to between 10 and 0 copies per reaction, subjected to real-time PCR with 12 replicates for each dilution step. Probit analysis (plot of fitted model) was performed [32] using StatGraphics Centurion XVI.I (16.1.11; Statgraphics Technologies, The Plains, Virginia, USA) to determine the LoD by fitting template copies against the cumulative fractions of positive PCR observations and used for calculating the lower and upper 95% confidence limits. The LoD of the 16S rRNA SNP RT-PCR was determined likewise using samples with 0-15 rRNA copies per reaction (12 replicates for each dilution step).

## 3. Results

### 3.1. Set-up and optimization of a new 16S rRNA gene allele-specific PCR assay

The “16S SNP BA probe” for hybridization to the *B. anthracis* specific sequence variation in 16S-BA-alleles in the *B. anthracis* genome was designed such as that the SNP position was located centrally. In order to increase fidelity of this probe, six locked nucleic acid (LNA) bases were introduced (Table 1). Likewise, the alternative “16S SNP BC probe” recognizing the non-*B. anthracis* specific 16S-BC-alleles of *B. anthracis* features five LNA positions (Table 1). The 16S SNP BA probe was verified *in silico* against the NCBI database to be highly specific for *B. anthracis*, only genomes of a few bacterial isolates exhibited identical sequences among these was e.g., a small number of *Sphingomonas* spp. Others, such as a few genomes annotated as *Staphylococus aureus* had the same one-base-pair mismatch at the SNP-position (relative to *B. anthracis*) and were thus identical to other *B. cereus s. l*. genomes, hybridizing perfectly against the alternative “16S SNP BC probe” (supplementary Figure S1).

Initially, the 16S SNP BC probe which deviates only by the one central SNP base from the 16S SNP BA probe also carried a fluorescent dye/quencher pair. However, since this probe was found to be not entirely specific for recognizing 16S rRNA fragments of *B. cereus s. l*. members, we decided to additionally design this SNP-competing probe as a fluorescently “dark” probe in order to reduce costs of synthesis (Table 1). Thus, the 16S rRNA SNP-PCR may be considered a pseudo-duplex assay (see below for details). All PCR runs were performed with both probes, typically with the 6FAM-labeled 16S SNP BA probe and the dark 16S SNP BC probe.

*In silico* analysis against the NCBI nt database confirmed that the PCR amplification primers 16S SNP F and 16S SNP R (Table 1) were not species-specific for *B. anthracis*. Indeed, besides DNA from other members of the *B. cereus s. l*. group, these primers would also amplify genome-sequences of various other bacteria, such as *Paenibacillus* spp., or the reverse primer would bind to sequences of *Alkalihalobacillus clausii* or *Bacillus licheniformis* among others. This ambiguity is not surprising for primers hybridizing against 16S rRNA gene sequences. Conversely, the pivotal factor for the detection assay introduced here is that only the 16S SNP BA probe hybridizes without any mismatch against 16S-BA-allele in *B*. *anthracis* (supplementary Fig. S1). Thus, the specificity of the PCR assay is uniquely and entirely governed by the LNA-enhanced 16S SNP BA probe.

The 16S rRNA SNP-PCR was robust for deviations from the optimum annealing temperature (62°C; supplementary Table S1). Also, primer (supplementary Table S2), probe (supplementary Table S3) and MgCl_2_ (supplementary Table S4) concentrations and pipetting errors (supplementary Table S5) were tolerated quite well. Intra- and inter-assay (supplementary Table S6 and S7) variability was determined with positive, weakly positive and negative template DNA. The average PCR variations were at 0.0-1.1% (intra-assay) and 1.1-1.2% (inter-assay), respectively (supplementary Table S6 and S7), indicating high precision of the PCR. Melt point analysis of the 16S-BA-allele PCR product vs. the 16S-BC-allele PCR product (supplementary Fig. S2) indicated specific amplification of each allele fragment.

### 3.2. Competitive amplification - inhibition of the 16S-BA-allele fragment-PCR by excess of the alternative 16S-BC-allele

Though the new 16S rRNA SNP-PCR assay was tested very robust and precise, we were wondering to which degree the assay would be inhibited by large excesses of the alternative 16S-BC-allele fragment featuring a single mismatch at the SNP located centrally in the hybridizing 16S-BA-allele specific PCR probe (supplementary Fig. S1). For testing this, we first evaluated which probe ratio (16S rRNA SNP BA vs. BC probe) would yield the lowest residual fluorescence values (in the 6FAM-channel of the 16S-BA-allele-specific probe) when providing only 16S-BC-allele containing DNA as PCR template. In these tests, the concentration of the 16S-BA-allele specific probe was kept constant at 0.25 µM. The resulting 6FAM-fluorescence values were very low compared to regular amplification (supplementary Table S8), signals were weakly linearly increasing and no Ct values were detected. The lowest fluorescence, barely above the negative control level, was recorded at a ratio of 0.25 / 0.75 µM (16S rRNA SNP-BA probe /-BC probe). Thus, this ratio was used for all following tests.

Next, a constant 100 template copies of the 16S-BA–allele fragment per reaction were titrated against increasing copy numbers of the alternative 16S-BC–allele fragment. Supplementary Figure S3 and supplementary Table S9 show that an excess of 16S-BC-allele to BA-allele fragments of 10^6^, 10^5^, 10^4^ or 10^3^ to 1 (supplementary Table S9; assay #1-4) inhibits detection of the 16S-BA-allele fragment. This is because there was neither any *bone fide* sigmoidal PCR amplification, nor were there any fluorescence signals with values meaningfully above the 16S-BC-allele-only controls (assays #11 and #12). Starting with 7.5 × 10^4^ copies of competing 16S-BC-alleles (vs. 100 16S-BA-allele copies, i.e., 750:1; assay #5) both a regular Ct value was provided and fluorescence started to markedly increase above base level. At a ratio of 500 to 1 (16S-BC- to BA-alleles), *B. anthracis* detection became possible (assays #6 vs. #12; #7). Latest at a surplus of equal or less than 100:1 (assay #8) detection of 16S-BA-allele among BC-alleles was robustly possible. Thus, at the very least a single copy of 16S-BA-allele can be detected in the presence of 100 BC-alleles.

### 3.3. Sensitivity and specificity of the 16S rRNA SNP-PCR assay

Similar to earlier work [33], we sought to harness the specificity of SNP-interrogation without assaying the alternative SNP state (i.e., the 16S-BC-allele here). Because detecting the 16S-BC-allele was not of interest for the assay at hand, the respective labelled 16S SNP BC probe was replaced by an unlabeled, fluorescently “dark” probe (i.e., a BA allele SNP-competitor probe; Table 1). In effect, primers would still amplify both alleles; however, the fluorescent probe for the 16S–BA-allele would be outcompeted by the dark probe on 16S-BC-allele targets and the fluorescent 16S rRNA BA SNP probe would only generate signals in the presense of cognate 16S-BA-allele sequences. Thus, this approach using a dark competing probe would diminish the inadvertent generation of unspecific fluorescence generated by mishybridization of 16S rRNA BA SNP probes to 16S-BC-allele sequences.

To formally validate the sensitivity of the 16S rRNA SNP-PCR assay, a panel of 14 different *B. anthracis* DNAs was employed. These *B. anthracis* strains represent all major branches A, B and C [34] including prominent sub-branches [35] of the global *B. anthracis* phylogeny (supplementary Table S10). All DNAs produced positive PCR results. Similarly, we tested a “specificity panel” of potentially cross-reacting organisms (supplementary Table S11). This panel included 13 DNAs of non-*anthracis B. cereus s. l*. strains. Also included were DNAs of common animal host organisms such as cattle, goat, sheep and human. Neither of these DNAs yielded any positive PCR results. Finally, DNAs of organisms relevant for differential diagnostics and other prominent microbial pathogens were also assayed by the new *B. anthracis* specific 16S rRNA SNP-PCR (supplementary Table S12). Again, none of these DNAs resulted in false-positive PCR results. Of note, *Sphingomonas zeae* JM-791 [36] harboring 16S rRNA genes 100% identical in the region of the 16S SNP BA probe but differences in the primer binding sites, yielded negative PCR results. These results clearly indicated that the new PCR is both sensitive and specific for *B. anthracis*.

### 3.4. Linear dynamic range, efficiency and limit of detection of the B. anthracis specific 16S rRNA SNP-PCR assay

The linear dynamic range of the new PCR was determined based on measurements of serial DNA dilutions using recombinant 16S-BA–allele fragments or genomic DNA of *B. anthracis* Ames, respectively, as templates (Figure 1). Linearity was observed over a range from 10^1^ to 10^9^ copies per reaction for cloned template DNA (Figure 1A; supplementary Table S13). In nine out of nine PCR replicates, positive signals were obtained down to 10^1^ copies per reaction. At 10^0^, two out of nine reactions were negative, thus defining the lower limit of the linear dynamic range. The coefficient of determination (R^2^) was calculated as >0.999. From the slope of the linear regression, the efficacy of the PCR was derived as 2.0 (which is 100.1% of the theoretical optimum). Thus, the 16S rRNA SNP-PCR assay performed very well over a wide 9 log_10_ concentration range of template DNA.

**Figure 1:**
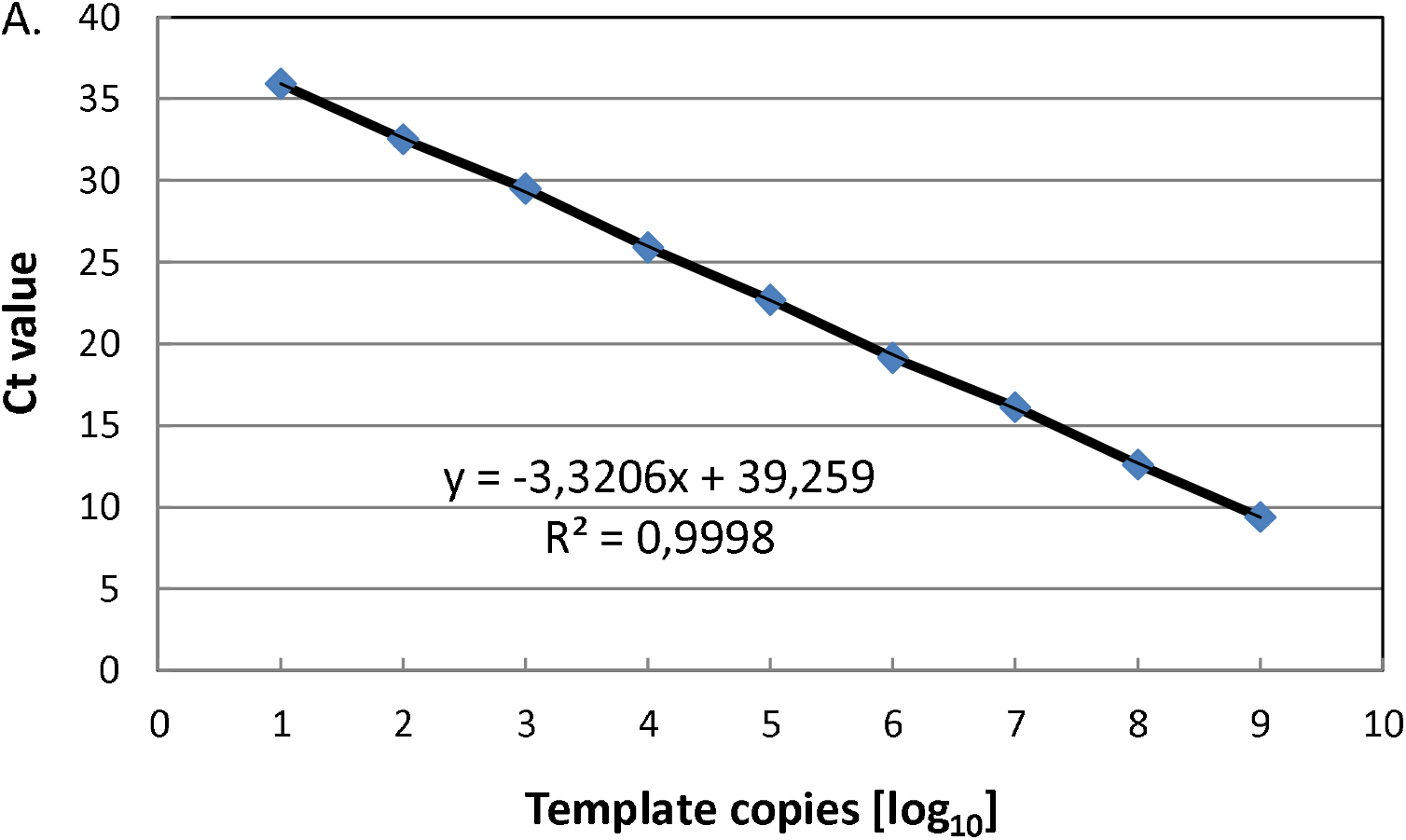
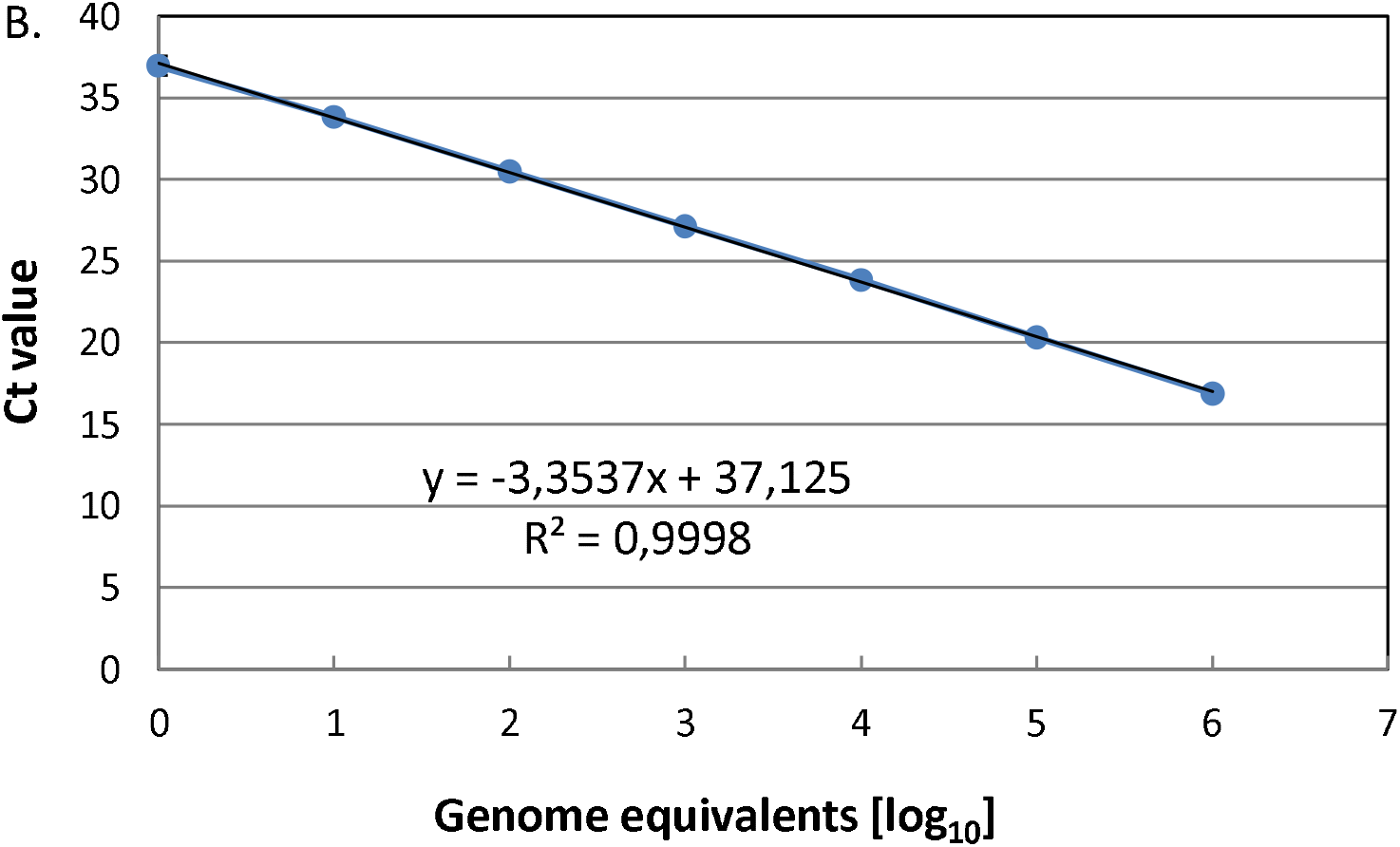
Linearity of the 16S rRNA SNP-PCR. Serial dilutions of DNA of (A.) a fragment comprising the 16S-BA-allele or (B.) *B. anthracis* strain Ames were serially diluted 1:10, PCR-tested and template copies (A.) or genome equivalents (B.) plotted against Ct values. Indicated in the graphs are the slopes of the linear regressions and the coefficients of determination (R^2^). Individual data points represent average values from n = 3 × 3 PCR-tests.

The *B. anthracis* Ames genome harbors four copies of the 16S-BA-allele and seven copies of the BC-allele. Linear range parameters were very similar to that of cloned 16S-BA-allele DNA-fragment (Figure 1B; supplementary Table S13). Because of the upper concentration limit of our *B. anthracis* Ames DNA preparations, the highest value in the linear range was 10^6^ genome copies. Thus, here the linear range covered target concentrations from 10^0^ to 10^6^ copies per reaction. The coefficients of determination (R^2^) was determined as >0.999 and the efficacy of the PCR as 1.99 (which is 98.7% of the theoretical optimum). This indicated that the 16S rRNA SNP PCR assay yielded very similar results in these experiments whether recombinant target DNA or authentic *B. anthracis* DNA was used as templates. Note though, a single *B. anthracis* Ames genome carries four copies of the 16S-BA-allele. This explains why all PCRs yielded positive signals with DNA template at 10^0^ copies (genome equivalents), whereas PCRs using single copy recombinant template did not.

Next, we determined the LoD for the 16S rRNA SNP-PCR assay by probit analysis (Figure 2; numerical data in supplementary Table S14). The assay had a limit of detection of 2.9 copies per reaction. This calculates to about 0.6 copies/µl with a probability of success of 95% with confidence interval of 2.4 - 4.5 copies/assay.

**Figure 2:**
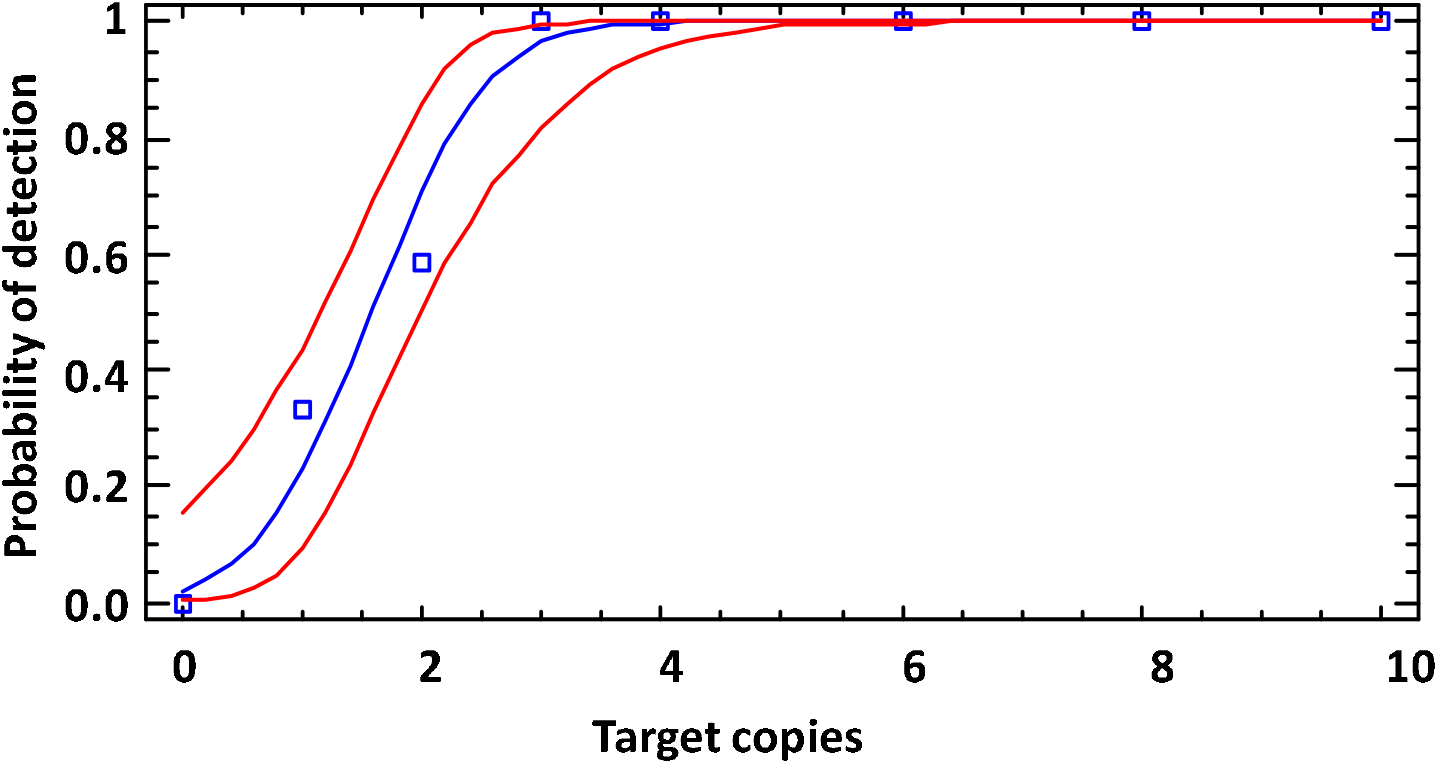
Limit of detection (LoD) of the 16S rRNA SNP-PCR (analytical sensitivity). DNA fragments comprising the 16S-BA-allele were diluted to the indicated copies per reaction (numerical data in supplementary Table S14) and subjected to real time PCR (12 replicates for each data point). Probit analysis (plot of fitted model) was performed to determine the LoD by fitting template copies against the cumulative fractions of positive PCR observations (blue squares and line) and used for calculating the lower and upper 95% confidence limits (red lines).

### 3.5. Comparison of the new 16S rRNA SNP-PCR assay with existing PCR assays

In order to further assess the performance of the 16S rRNA SNP-PCR assay, we compared it with other established PCR assays for *B. anthracis* identification currently used in our laboratory. These assays target the single copy genes *dhp61* [9] or *PL3* [6] that have been individually validated before and compared to other commonly used *B. anthracis* PCRs [11]. Using log_10_ dilutions of *B. anthracis* Ames DNA, the 16S rRNA SNP-PCR exhibited markedly, at least three units, lower Ct values (27.9±0.4; 31.7±0.1; 35.4±0.7) than *dhp61* (32.1±0.0; 35.4±0.6; 38.9±1.5) or *PL3* (31.8±0.2; 36.1±0.7; >40) at 1,000, 100 or 10 genome equivalents, respectively (Fig. 3). *B. cereus* DNA did not result in amplification by any PCR assay. This result strongly suggested that the multi-copy 16S rRNA SNP-PCR assay performs competitively when compared back-to-back with established PCR assays for the detection of *B. anthracis*.

**Figure 3:**
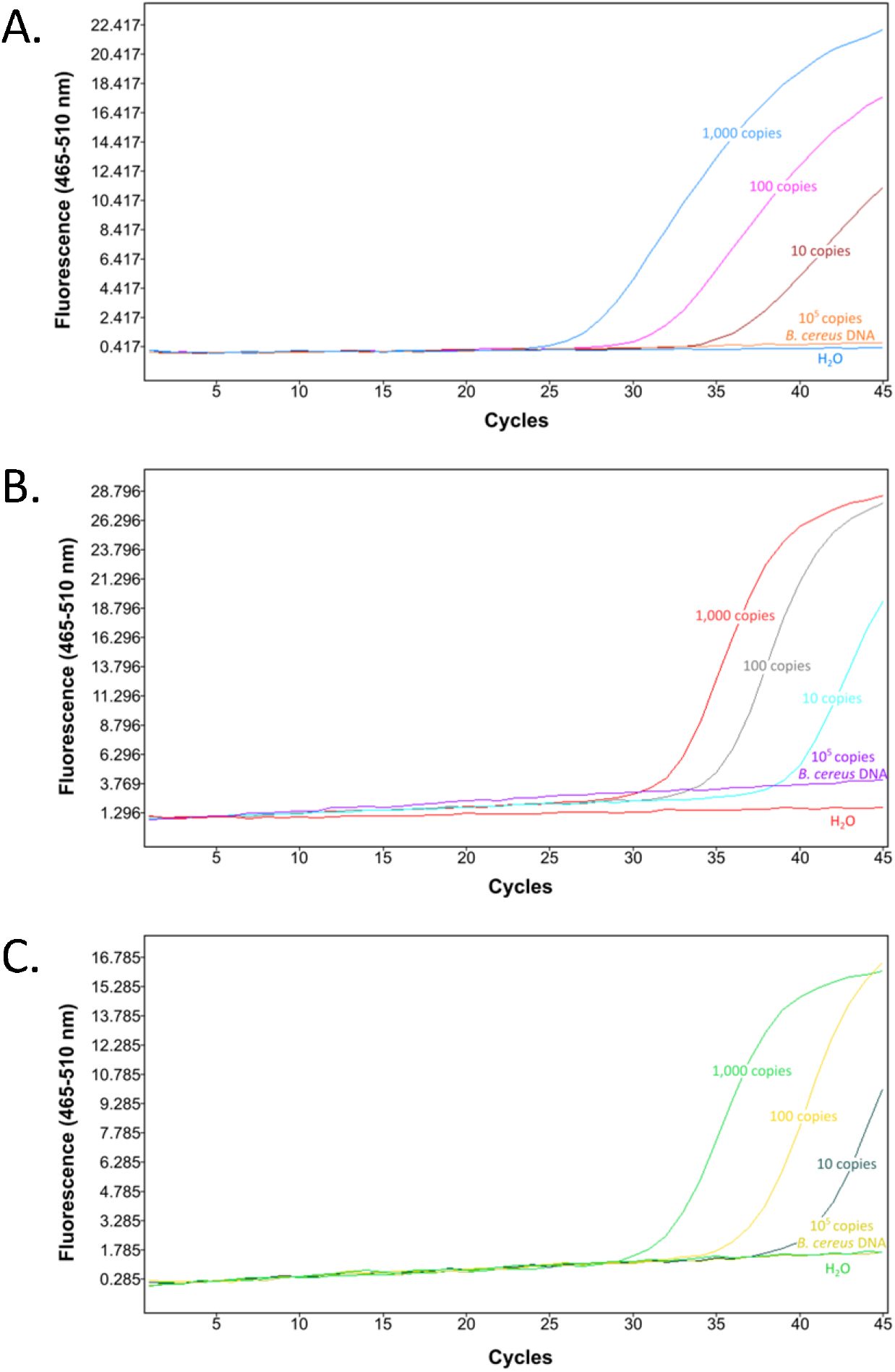
Comparison of the new 16S rRNA SNP-PCR assay with existing PCR assays. Different numbers of *B. anthracis* Ames template DNA (1,000, 100 or 10 genome equivalents per reaction), non-target DNA (10^5^ templates of *B. cereus* DNA) or water (negative) control were subjected to real time PCR using the new 16S rRNA SNP assay (A), published *dhp61* gene assay [9] (B) or published *PL3* gene assay [6] (C). Representative amplification curves (from n=3 with similar results) are shown.

### 3.6. Challenge of the new 16S rRNA SNP-PCR assay with samples from a ring trial

Along this line of reasoning, we next challenged the 16S rRNA SNP-PCR assay with samples from a previous ring trial for *B. anthracis* nucleic acid detection [27]. Again, the test was performed in comparison with the established PCR assays for *B. anthracis* identification, *dhp61* [9] and *PL3* [6]. Each of the assays was able to correctly identify the two positive out of four samples (supplementary Figure S4). Similar to evaluating known concentrations (Figure 3), the 16S rRNA SNP-PCR assay performed the best. It yielded the lowest Ct values (supplementary Figure S4), at least 2 units lower than that of *dhp61* or *PL3* PCR. The 16S rRNA SNP-PCR assay may thus be ideally suited for this kind of analysis in which low target DNA quantities can be expected.

### 3.7. Challenge of the new 16S rRNA SNP-PCR assay with total DNA from spiked soil samples

Since the 16S rRNA SNP-PCR assay performed well thus far even in the presence of *E. coli* and human (supplementary Figure S4) or competing *B. cereus* (supplementary Figure S3) DNA, we evaluated to what extent the assay would be able to detect target DNA in spiked soil samples. These samples were spiked with cells of *E. coli* and *F. tularensis* and cells or endospores of *B. anthracis* and/or *B. thuringiensis* and were subjected to DNA purification. As above, the 16S rRNA SNP-PCR assay was conducted in comparison with the established PCR assays for *B. anthracis* identification *dhp61* [9] and *PL3* [6]. Supplementary Figure S5 shows the PCR amplification curves. Samples #1, #2 and #4 were samples spiked with *B. anthracis*, sample #3 only contained *E. coli* and *B. thuringiensis*. Sample #4 had a large excess of *B. thuringiensis* over *B. anthracis* (a factor of 10^4^). The 16S rRNA SNP-PCR assay detected *B. anthracis* in samples #1 and #2 but not in #4. Conversely, *dhp61* or *PL3* assays detected all three positive samples. The failure to detect *B. anthracis* by the 16S rRNA SNP-PCR assay in sample #4 is in line with our initial tests using massive excess of *B. cereus* DNA competing with *B. anthracis* detection (supplementary Figure S3; Table S9). Notably, the 16S rRNA SNP-PCR exhibited markedly, about three units, lower Ct values (23.6±0.7 or 16.4±0.0) than *dhp61* (26.2±0.1 or 19.9±0.1) or *PL3* (25.7±0.0 or 19.5±0.1) for samples #1 and #2, respectively. This result confirmed our preceding findings that the 16S rRNA SNP-PCR assay can reach a lower detection limit than established assay as long as there is no large excess of other *B. cereus s*.*l*. DNA competing for amplification primers.

### 3.8. The new 16S rRNA SNP-PCR assay also functions as a RT-PCR assay

We reasoned that the real time 16S rRNA SNP-PCR assay targeting *B. anthracis* DNA may be converted into a RT-PCR assay targeting RNA in the form of 16S-BA-allele transcripts that harbor the *B. anthracis*-specific SNP. In order to test this, cells of *B. anthracis* Sterne or *B. cereus* 10987 were grown to exponential growth phase, inactivated and total nucleic acids (including genomic DNA) was isolated alongside parallel preparations of DNA only. The one-step RT-PCR reaction was thus run with a mixture of genomic DNA and RNA, which can both be targeted by the assay. For comparison, the above validated 16S rRNA real time SNP-PCR was conducted in parallel with genomic DNA as the only template (no RT-reaction). When using identical samples, RT-PCR reactions (with templates consisting of total RNA and DNA) resulted in intensely lower Ct values than without reverse transcription (since only genomic DNA served as template; Fig. 4). This result indicated that the 16S rRNA SNP-PCR assay functions both for DNA- and RNA-based (RT) PCR. Notably, differences in Ct values (RT-PCR vs. PCR) were in the range between 9 and 10 units. This translates to an about 1,000-fold improvement using RT-PCR over DNA-only PCR.

**Figure 4:**
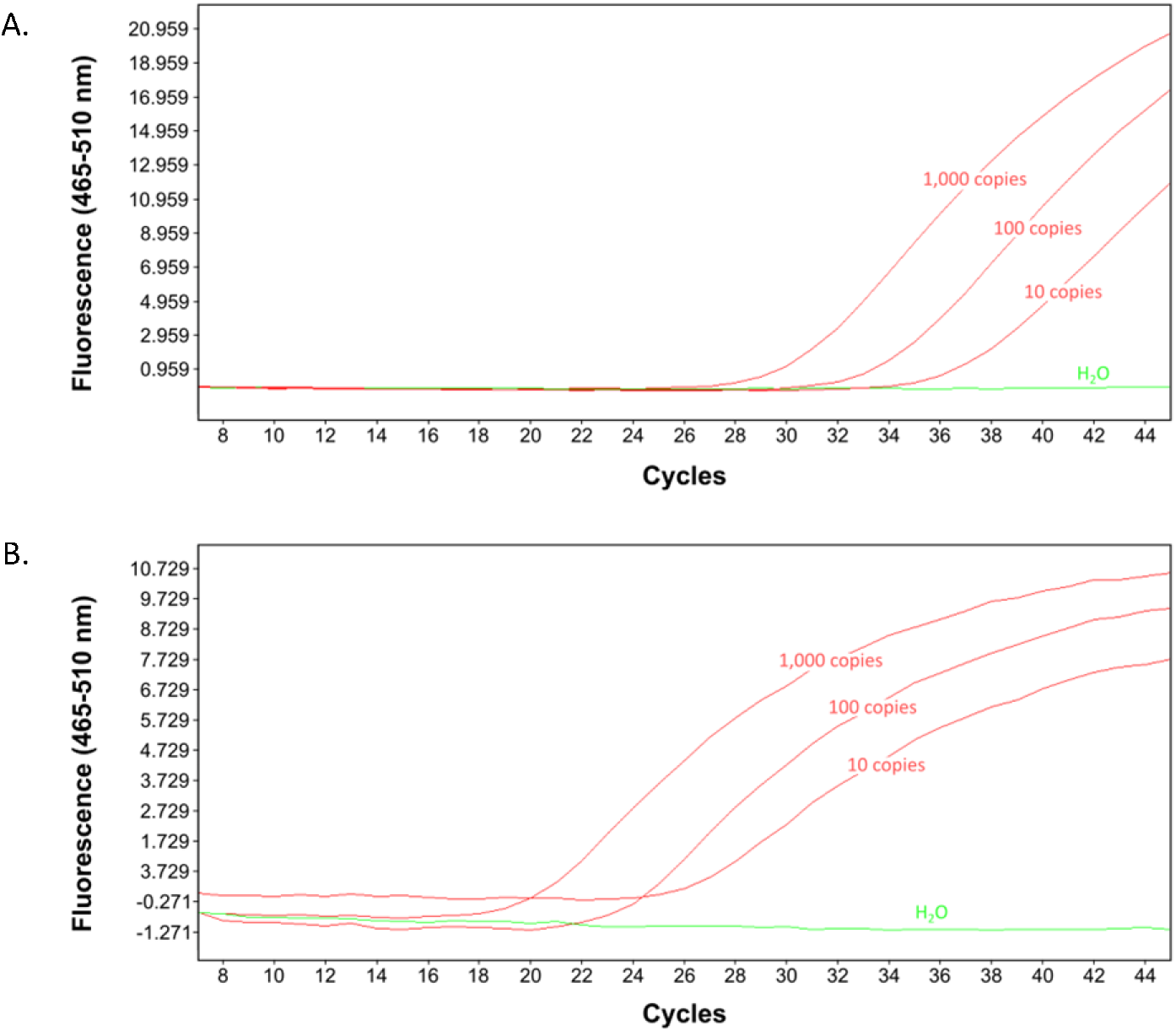
Comparison of the 16S rRNA SNP-PCR assay (DNA-only) with the RT-16S rRNA SNP-PCR assay (DNA+RNA). Total DNA or total DNA+RNA isolated from exponentially growing cells of *B. anthracis* or *B. cereus*, respectively, were used for PCR amplification of 16S-BA-allele DNA (A) or additionally after reverse transcription of 16S rRNA (ribosomal RNA) (B). Representative amplification curves (from n=3 with similar results) are shown.

**Figure 5:**
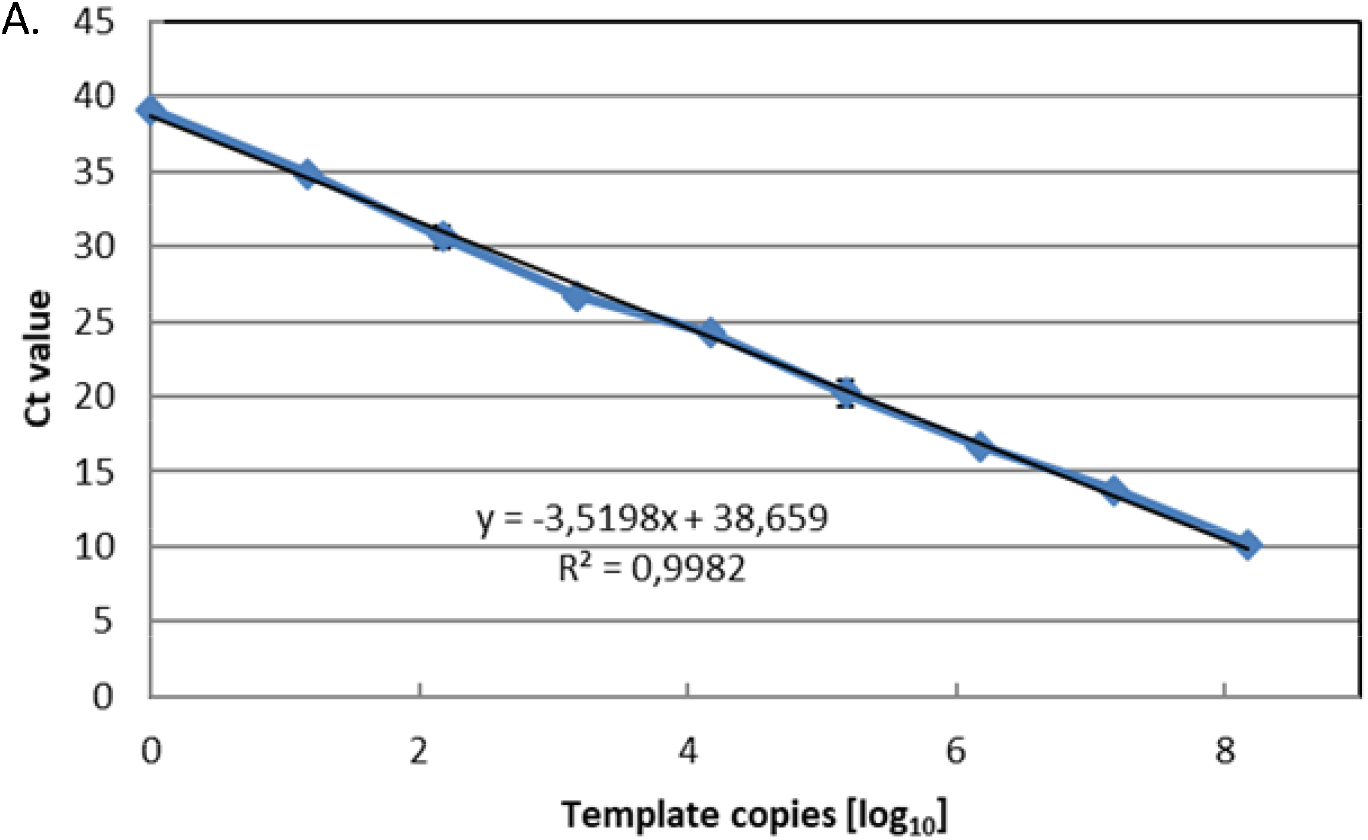
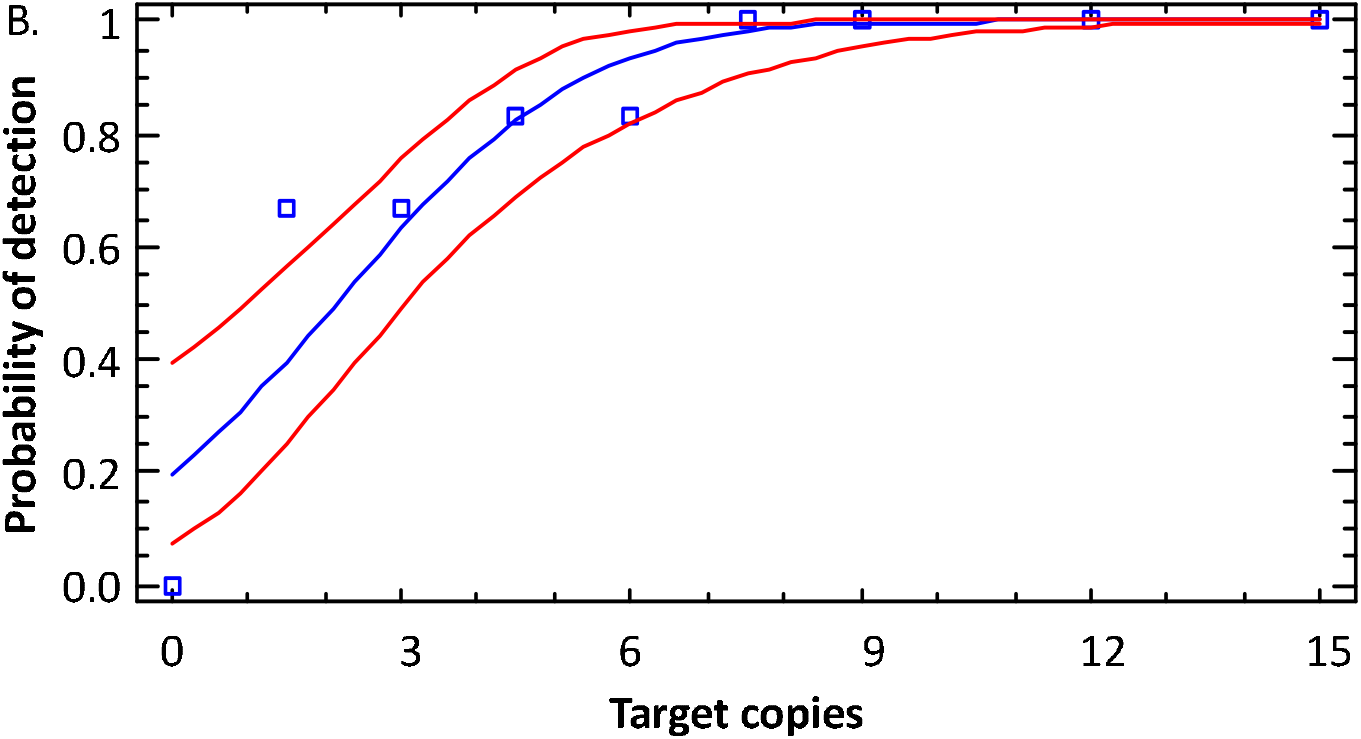
Linearity and LoD of the 16S rRNA SNP RT-PCR. Serial dilutions of RNA (with DNA) of *B. anthracis* strain Sterne were serially diluted 1:10, RT-PCR-tested and template copies plotted against Ct values (A). Indicated in the graph is the slope of the linear regression and the coefficients of determination (R^2^). Individual data points represent average values from n = 3 × 3 PCR-tests. Analytical sensitivity of the 16S rRNA SNP RT-PCR was determined by diluting samples from (A) to the indicated copies per reaction (numerical data in supplementary Table S16) and subjected to RT-PCR (12 replicates for each data point). To determine the LoD, probit analysis (plot of fitted model, blue squares and line) was performed (as in Fig. 2) and the lower and upper 95% confidence limits (red lines) determined (B).

### 3.9. Linear dynamic range, efficiency and limit of detection of the B. anthracis 16S rRNA SNP RT-PCR assay

To further characterize the RT-PCR, we determined the linear dynamic range and determined the LoD (Probit) of the 16S rRNA SNP RT-PCR using total RNA/DNA of *B. anthracis* Sterne (similar to DNA-only templates, see above). The RT-PCR was linear over a range from 10^0^ to 10^8^ template rRNA+DNA per reaction (Figure 4A; supplementary Table S15). The coefficient of determination (R^2^) was 0.9982 and the efficacy of the RT-PCR was 1.92 (which is 92.3% of the theoretical optimum). Thus, the 16S rRNA SNP RT-PCR assay performed well over a wide 9 log_10_ concentration range of template RNA+DNA (higher template numbers than 1.5 × 10^8^ were not tested).

The LoD for the 16S rRNA SNP RT-PCR assay as determined by probit analysis (numerical data in supplementary Table S16) was 6.3 copies per reaction. This calculates to about 1.3 copies/µl with a probability of success of 95% with confidence interval of 5.0 – 8.9 copies/assay. Thus, the RT-PCR reaction performed similarly well as the PCR reaction.Mindful of the about 3 log_10_ units higher number of 16S rRNAs in cells than genomes, detection of *B. anthracis* with the rRNA-directed RT-PCR is superior to the respective real time PCR assay and all other *B. anthracis* PCR assays tested.

## 4. Discussion

Use of SNPs as reliable markers for the identification of *B. anthracis* among its closest relatives of the *B. cereus* group is not a novel approach. This has previously been achieved with high specificity and sensitivity for nucleotide position 640 in the *plcR* gene [10] or at position 1050 in the *purA* gene [33] and diverse assays were thoroughly evaluated in [11]. Likewise, ribosomal gene sequences and intergenic transcribed spacers (ITS) between 16S and 23S rRNA genes have also been employed for *B. anthracis* identification in the past [37-40]. However, while these authors focused on the specific identification of *B. anthracis*, they neglected the potential of developing a sensitive assay making use of the multi-copy nature of their targets. An interesting exception is a study on fluorescent DNA-heteroduplex detection of *B. anthracis* [41]. Herein detection was preceded by general PCR-amplification of a fragment of the 16S rRNA gene region of *B. cereus s. l*. group strains containing a presumably specific SNP (pos. 980). This SNP, however, is neither specific for *B. anthracis* nor for the *B. cereus s*.*l*. group [24]. Anyway, Merrill et al. succeeded in establishing a LoD for their PCR of approximately 0.05 pg of purified *B. anthracis* genomic DNA (which can be calculated to represent 10 to 20 cell equivalents per reaction) [41]. This is higher than the LoD of about 1 to 2 cell equivalents per reaction found in our study. More importantly, Merrill et al. also took the effort to determine the detection limit of their presumably specific SNP in mixtures of 16S rRNA gene amplicons from *B. anthracis* and *B. cereus* [41]. The authors observed a detection limit of 1 out of 50 for *B. anthracis* DNA mixed with *B. cereus* DNA. They explained this limit as narrowed by methodological constraints and from competitive hybridization dynamics during probe annealing [41]. This finding can be compared with our results. The PCR assay developed here was able to detect at least one *B. anthracis*16S-BA-allele target among 100 BC-allele targets (supplementary Fig. S3 and supplementary Table S9). At higher alternative (16S-BC-allele) concentrations, these templates will outcompete the 16S-BA-allele for primer binding. Thus, the higher the fraction of 16S-BC-allele, the lower the relative amplification of 16S-BA-allele resulting in increasingly non-exponential amplification of the latter. In contrast, for a SNP in the DNA target *plcR* used for the differentiation of *B. anthracis* from *B. cereus*, a 20,000-fold excess of the alternate *B. cereus* allele did not preclude the detection of the *B. anthracis* allele [42]. With *B. cereus* spore counts in soils spanning a wide range of 1 × 10^1^ to 2.5 × 10^4^ CFU per g soil [43], the *plcR* SNP-PCR should be able to detect *B. anthracis* in practically any sample. Here, the new 16S rRNA SNP-PCR on DNA as target molecule would fall short with only covering up to medium *B. cereus*-loaded soils. However, when targeting ribosomal RNA the sensitivity (LoD) of the 16S rRNA SNP RT-PCR would be at least three orders of magnitude increased. Then it should be possible to challenge the LoD values achieved by the *plcR* SNP-PCR (25 fg DNA or about 5 genome equivalents) [42].

A potential limitation of the multi-copy nature of the 16S-BA-allele may be the variable abundance of this allele in different *B. anthracis* strains. Previously, we could show that most *B. anthracis* strains harbor 3 (58.39%) 16S-BA-alleles. There are, however also a number of isolates only possessing 2 (23.04%), 4 (17.10%) 5 (1.15%) and a single one with only 1 (0.31%) 16S-BA-alleles [25]. Thus, in most cases this multi-copy gene allele can be harnessed nevertheless. A more typical multi-copy marker for detection of bacterial biothreat agents (and of other pathogens) constitute insertion sequence (IS) elements, which are wide-spread mobile genetic entities. For instance in *Brucella* spp. *IS711* occurs in multiple genomic copies and thus, the detection of this *IS711* is very sensitive. *B. melitensis* and *B. suis* contain seven complete copies, *B. abortus* carries six complete and one truncated *IS711* copies, *B. ovis, B. ceti* and *B. pinnipedialis* even more than 20 copies [44]. Consequently, the lowest concentration of *Brucella* sp. DNA that could be detected was about ten times lower for *IS711* than e.g., for single copy genes *bcsp31* (*Brucella* cell surface 31 kDa protein) or *per* (perosamine synthetase), respectively [45]. Similarly, in *Coxiella burnetii*, detection sensitivity of specific *IS1111* was compared to that of the single-copy *icd* gene (isocitrate dehydrogenase) [46]. While both PCRs for *icd* and *IS1111* had similar LoDs of 10.38 and 6.51, respectively, sensitivity of *IS1111* was still superior because of its multiple copy nature. Between 7 and 110 copies of this mobile element were found in various *C. burnetii* isolates [46].

The differences in threshold values (ΔCt = 9.96±0.65) of identical samples obtained from (RT)-PCR using 16S-BA-allele DNA-only vs. 16S-BA-alleleDNA+RNA is enormous. There is an approximate factor of about 1,000 (2^9.96^) times more template in the DNA+RNA sample than in the DNA-only sample. This factor favorably agrees with the numbers of genome copies and 16S rRNA transcripts in cells [17, 18]. Similar to the work at hand, earlier work employed a combination of a DNA multi-copy marker and sensitive detection of rRNA transcript targets in *Mycobacterium ulcerans* [20]. The authors determined a LoD of 6 copies of the 16S rRNA transcript target sequence. For comparison, a LoD of two target copies of the high-copy insertion sequence element *IS2404* which is present in 50 to 100 copies in different *M. ulcerans* strains was calculated from parallel experiments [20]. Ribosomal RNA detection was also utilized for *Mycobacterium leprae* diagnosis by the same research team. Here, a LoD of three *M. leprae* target copies was achieved for a novel 16S rRNA RT-PCR assay; the same value as determined for the *M. leprae* specific multi-copy repetitive DNA target assayed in parallel [21]. On first glance these values do not especially speak in favor of querying for 16S rRNA transcripts, however, one has to consider the high numbers of these molecules per cell in comparison to DNA markers (including the high-copy ones). Thus, the chance of capturing one of the more abundant rRNA molecules should be higher than that of the more limited DNA molecules. Indeed, this idea was explored e.g., for *Escherichia coli, Enterococcus faecalis, Staphylococcus aureus, Clostridium perfringens*, and *Pseudomonas aeruginosa* by [19]. Comparative quantitative detection of these bacteria by RT-PCR (16S rRNA) vs. PCR (16S rRNA genes) revealed that the rRNA-detecting assay was 64- to 1,024-fold more sensitive than the one detecting DNA. Similarly, work on pathogenic spirochete *Leptospira* spp. found that 16S rRNA-based assays were at least 100-fold more sensitive than a DNA-based approach [47]. These authors also found that Leptospiral 16S rRNA molecules remain appreciably stable in blood. From this insight, the authors then highlighted the potential use of 16S RNA targets for diagnosis of early infection. Nevertheless, potential limitations of this approach were also noted. Efficacy of the required reverse transcription reaction has to be considered, RNA molecules are notoriously less stable than other biomarkers and their cellular abundance (and as a consequence their detection) can be expected to be variable [47]. Finally, though for qualitative detection not required, absolute quantification of microbial cells based solely on enumeration of RNA molecules is complicated because of these variations in transcript numbers depending, e.g. on growth phase [47]. However, the cell numbers determined by RT-PCR were similar when compared along-side standard methods such as cell counts, PCR or fluorescence in situ hybridization (FISH) [48]. Yet, in certain instances, there might be an additional advantage of performing PCR on rRNA directly (via RT-PCR) instead of targeting DNA (including DNA of rRNA genes). Because DNA is more stable than RNA, DNA may originate from both live and dead bacterial cells. In contrast, rRNA molecules may be considered to be more closely associated with viable bacteria [49]. Though this might also be possible with the new PCR assays introduced in the work at hand, we chose to combine DNA and rRNA detection in a single test-tube for the sake of simplicity (no troublesome DNase treatment of purified RNA required) and depth of detection.

## 5. Conclusions

In this work, we designed and validated a new PCR-based detection assay for the biothreat agent *B. anthracis*. This assay can be run as a real-time PCR with solely DNA as template or as a RT-real-time version using both cellular nucleic acid pools (DNA and RNA) as template. This assay was found to be highly species specific yielding no false positives and was sensitive with a LoD of about 0.6 copies/µl (DNA-only) and about 1.3 copies/µl (DNA+RNA). With the high abundance of 16S rRNA moieties in cells this assay can be expected to facilitate the detection of *B. anthracis* by PCR.

## Data Availability

All relevant data is included in the manuscript and its supplementary file.

## Supplementary Materials

The following are available online: Supplementary Tables 1 – 16; Supplementary Figures 1 – 6.

## Author Contributions

Conceptualization, G.G. and P.B; investigation, P.B., M.D.T.N., M.C.W.; methodology, M.D.T.N., and P.B.; formal analysis and validation, P.B., M.D.T.N. and G.G.; resources, G.G. and M.C.W.; data curation, P.B., M.D.T.N. and M.C.W.; writing— original draft preparation, G.G., and P.B.; writing—review and editing, P.B., M.D.T.N., M.C.W. and G.G.; visualization, M.D.T.N., P.B. and G.G.; supervision and project administration, G.G., and P.B.; funding acquisition, G.G. All authors have read and agreed to the published version of the manuscript.

## Acknowledgments

The authors thank Mandy Knüper (Bundeswehr Institute of Microbiology) for support with probit-analysis and the gift of DNA from spiked soil samples, Stefanie Gläser (Justus-Liebig-University Gießen, Germany) Erwin Märtlbauer (Ludwig-Maximilians-University, Munich, Germany) and Monika-Ehling Schultz (University of Veterinary Medicine, Vienna, Austria) for several *B. cereus* strains, as well as Paul Keim (Northern Arizona University, Flagstaff, USA), and Wolfgang Beyer (Hohenheim University, Stuttgart, Germany) for the gift of *B. anthracis* strains/DNA. Thanks are due to Rahime Terzioglu for technical assistance and Olfert Landt (TIB Molbiol, Berlin) for support in probe design.

