## Supplementary Material for "Ultrasensitive detection of *Bacillus anthracis* by real time PCR targeting a polymorphism in multi-copy 16S rRNA genes and their transcripts"

### 6. Supplementary Material

#### *6.1. Supplementary Tables*

Supplementary Table S1: Temperature optimum.

| Annealing temperature (°C) | Ct-values (Ø±SD)* | Fluorescence (arbitrary units)** |
|---|---|---|
| 61 | 33.1±0.1 | 27.9±0.9 |
| 62 | 32.7±0.2 | 32.1±1.1 |
| 63 | 32.5±0.2 | 28.0±0.9 |
| 64 | 32.7±0.1 | 25.2±0.4 |

* averages (Ø) ± standard deviations (SD) of n = 3 tests; ** value at the end of the 42th PCR cycle.

Supplementary Table S2: Primer titration.

| Primer concentrations (µM) | Ct-values (Ø±SD)* | Fluorescence (arbitrary units)** |
|---|---|---|
| 0.5 /0.5 | 26.6±0.9 | 13.8±0.4 |
| 0.25 / 0.5 | 26.5±0.5 | 9.7±0.4 |
| 0.5 / 0.25 | 27.2±0.1 | 11.7±0.2 |
| 0.25 / 0.25 | 26.6±0.1 | 6.0±0.4 |
| 1 / 0.5 | 27.6±0.2 | 12.8±0.6 |
| 0.5 / 1 | 27.3±0.2 | 15.0±0.4 |
| 1 / 1 | 26.6±0.7 | 13.0±0.5 |

* Average (Ø) ± standard deviations (SD) of n = 3 tests; ** Value at the end of the 42th PCR cycle.

Supplementary Table S3: Probe titration.

| Probe concentration BA* / BC** (µM) | Ct-values (Ø±SD)*** | Fluorescence (arbitrary units)**** |
|---|---|---|
| 0.25 / 0.25 | 27.5±0.3 | 15.9±0.2 |
| 0.125 / 0.25 | 27.5±0.2 | 9.2±0.5 |
| 0.25 / 0.125 | 27.8±0.1 | 16.2±0.1 |
| 0.125 / 0.125 | 27.4±0.6 | 9.9±0.5 |
| 0.5 / 0.25 | 27.3±0.6 | 24.8±1.6 |
| 0.25 / 0.5 | 27.5±0.3 | 14.7±0.3 |
| 0.5 / 0.5 | 27.7±0.4 | 23.2±1.7 |

* 6-FAM-labeled (hybridizes against the *B. anthracis*-specific allele of the 16S rRNA gene); ** „dark" probe lacking fluorescent label (hybridizes against the *B. cereus* allele of the 16S rRNA gene); *** averages (Ø) ± standard deviations (SD) of n = 3 tests; **** value at the end of the 42th PCR cycle.

Supplementary Table S4: Magnesium ion titration.

| $MgCl_2$ concentration (mM) | Ct-values (Ø±SD)* | Fluorescence (arbitrary units)** |
|---|---|---|
| 1 | 28.8±0.3 | 2.8±0.3 |
| 2 | 27.5±0.7 | 10.0±0.5 |
| 3 | 28.1±0.4 | 23.3±1.9 |
| 4 | 27.8±0.4 | 21.3±1.2 |

* averages (Ø) ± standard deviations (SD) of n = 3 tests; ** value at the end of the 42th PCR cycle.

Supplementary Table S5: Pipetting error (5%).

| Reaction volume (µl) | Ct-values (Ø±SD)* | Fluorescence (arbitrary units)** |
|---|---|---|
| 19 | 28.6±0.4 | 47.3±6.0 |
| 20 | 28.9±0.1 | 46.4±3.8 |
| 21 | 29.1±0.1 | 44.5±5.4 |

* averages (Ø) ± standard deviations (SD) of n = 3 tests; ** value at the end of the 42th PCR cycle.

Supplementary Table S6: Intra assay variability*.

| Strain | Assay 1 | Assay 2 | Assay 3 | Ct-values (Ø±SD)** | CV*** |
|---|---|---|---|---|---|
| L3-2641 | 32.4 | 32.7 | 33.1 | 32.7±0.3 | 1.0 |
| L3-2649 | 34.3 | 33.8 | 33.6 | 33.9±0.3 | 1.1 |
| L3-2657 | 33.8 | 33.9 | 33.9 | 33.9±0.1 | 0.2 |
| L3-2665 | 31.7 | 31.9 | 31.9 | 31.8±0.1 | 0.4 |
| L3-2866 | 31.7 | 31.9 | 31.9 | 31.8±0.1 | 0.4 |
| L3-3391 | 24.2 | 24.2 | 24.3 | 24.2±0.0 | 0.3 |
| L3-3399 | 22.8 | 22.7 | 22.7 | 22.7±0.0 | 0.1 |
| L3-3407 | 23.4 | 23.4 | 23.3 | 23.4±0.0 | 0.2 |
| L3-3408 | 22.9 | 22.9 | 22.9 | 22.9±0.0 | 0.3 |
| L3-3415 | 22.4 | 22.4 | 22.4 | 22.4±0.0 | 0.0 |
| L3-3416 | 23.5 | 23.6 | 23.7 | 23.6±0.1 | 0.3 |
| L3-3421 | 22.5 | 22.4 | 22.2 | 22.4±0.2 | 0.2 |

* replicates (n=3) of each PCR reaction with target DNA were run on the same day; ** averages (Ø) ± standard deviations (SD) of n = 3 tests; *** coefficient of variation (%).

Supplementary Table S7: Inter assay variability*.

| Template copies | Assays day 1 | Assays day 2 | Assays day 3 | Ct-value (Ø±SD)** | CV*** |
|---|---|---|---|---|---|
| $10^4$ | 25.9; 25.8; 26.0 | 26.2; 26.3; 26.3 | 25.6; 25.6; 25.6 | 25.9±0.3 | 1.2 |
| $10^2$ | 32.3; 32.9; 32.3 | 32.9; 33.0; 32.8 | 32.4; 32.2; 32.0 | 32.5±0.3 | 1.1 |

* replicates (n=3) of each PCR reaction with target DNA were run on three consecutive days; ** averages (Ø) ± standard deviations (SD) of n = 3 x 3 tests; *** coefficient of variation (%).

Supplementary Table S8: Probe titration in the presence of $10^9$ copies of 16S-BC-allele (absence of 16S-BA-allele).

| Probe concentration BA*/BC** (µM) [BA/BC] | Ct-values | Fluorescence (arbitrary units)**** |
|---|---|---|
| 0.25 / 0.25 [1:1] | -*** | 1.6±0.0 |
| 0.25 / 0.5 [1:2] | -*** | 1.1±0.4 |
| 0.25 / 0.75 [1:3] | -*** | 0.82±0.4 |
| 0.25 / 1.0 [1:4] | -*** | 1.4±0.3 |

* 6-FAM-labeled (hybridizes against the *B. anthracis*-specific 16S-BA-allele of the 16S rRNA gene); ** „dark" probe lacking fluorescent label (hybridizes against the *B. cereus* 16S-BC-allele of the 16S rRNA gene); *** no regular amplification, no CT-value; **** value at the end of the 42th PCR cycle.

Supplementary Table S9: Optimized probe concentration for the detection of the 16S-BA-allele (100 target copies) in the presence of increasing copies of the alternative BC-allele.

| Assay Number | 16S-BC-allele concentration (copies per reaction)* | Ct value (Ø±SD)** | Fluorescence (arbitrary units)*** |
|---|---|---|---|
| 1 | $10^8$ | **** | 3.63±0.3 |
| 2 | $10^7$ | **** | 3.44±0.2 |
| 3 | $10^6$ | **** | 3.35±0.1 |
| 4 | $10^5$ | 33.4±0.6 | 3.8±0.1 |
| 5 | 7.5 x $10^4$ | 34.0±1.3 | 4.8±0.7 |
| 6 | 5 x $10^4$ | 35.0±0.4 | 5.2±0.5 |
| 7 | 2.5 x $10^4$ | 34.2±0.3 | 6.9±0.3 |
| 8 | $10^4$ | 33.4±0.8 | 8.6±0.8 |
| 9 | $10^3$ | 32.6±0.1 | 17.9±0.6 |
| 10 | $10^2$ | 32.9±0.1 | 25.7±0.7 |
| 11 | no BA. only $10^5$ BC | **** | 2.6±0.1 |
| 12 | no BA. only 0.5 x $10^5$ BC | **** | 2.3±0.4 |
| 13 | positive control (only BA) | 33.0±0.2 | 31.3±4.3 |
| 14 | negative control ($H_2O$) | - | - |

* 16S-BA-allele concentration constant at 100 copies per reaction; ** averages (Ø) ± standard deviations (SD) of n = 3 tests; *** value at the end of the 42th PCR cycle; **** no regular amplification, no CT-value.

Supplementary Table S10: Sensitivity panel - target organism *B. anthracis*.

| # of *B. anthracis* strains | Phylogeny* | PCR result** |
|---|---|---|
| 1 | C.Br. A1055 | positive |
| 3 | B.Br. CNEVA | positive |
| 1 | B.Br. Kruger B | positive |
| 5 | A.Br. 001/002 | positive |
| 1 | A.Br. 011/009; A.Br.118 (STI) | positive |
| 1 | A.Br. 005/006 | positive |
| 1 | A.Br. Vollum | positive |
| 1 | A.Br. Aust 94 | positive |
| 5 | A.Br. Aust 94; A.Br.014 | positive |
| 1 | A.Br. Aust 94; A.Br.015 | positive |
| 2 | A.Br. 008/011; A.Br.127 (Pstr) | positive |
| 1 | A.Br. 008/011; A.Br.127(BUL) | positive |
| 1 | A.Br. 008/011; A.Br.161 (Heroin) | positive |
| 1 | A.Br. WNA | positive |

* phylogeny according to (1) and (2); ** results of n = 3 tests.

Supplementary Table S11: Specificity panel - potentially cross-reacting organisms.

| Organism | Strain number | PCR result** |
|---|---|---|
| *B. cereus* | ATCC 10987 | negative |
| *B. cereus* | 2998 | negative |
| *B. cereus* | 3093 | negative |
| *B. cereus* | LGL 3094 | negative |
| *B. cereus* | ATCC 4342 | negative |
| *B. cereus* bv. anthracis | CI-1 | negative |
| *B. cereus* bv. anthracis | CA-1 | negative |
| *B. cereus* | ATCC 33019 | negative |
| *B. thuringiensis* | ATCC 10792 | negative |
| *B. thuringiensis* | DSM 046 | negative |
| *B. paranthracis* | 2002 | negative |
| *B. weihenstephanensis* | B-0293 | negative |
| *B. mycoides* | B-298 | negative |
| *B. subtilis* | ATCC 6091 | negative |
| *B. megaterium* | ATCC 14581 | negative |
| *Homo sapiens**** | n.a.* | negative |
| *Bos taurus**** | n.a.* | negative |
| *Capra aegagrus hircus**** | n.a.* | negative |
| *Ovis gmelini aries**** | n.a.* | negative |
| *Equus caballus**** | n.a.* | negative |

* not applicable (n.a.); ** results of n = 3 tests; ***typical host organisms.

Supplementary Table S12: Specificity panel – organisms relevant for differential diagnostics and other pathogens.

| Organism | Strain number | PCR result* | Organism | Strain number | PCR result* |
|---|---|---|---|---|---|
| *Brucella sp.* | F-070660 | negative | *Moraxella catarrhalis* | B-0433 | negative |
| *Burkholderi mallei* | L3-2962 | negative | *Neisseria meningitidis* | B-1332 | negative |
| *Burkholderi pseudomallei* | L3-0711 | negative | *Propionibacterium acnes* | B-0438 | negative |
| *Burkholderia thailandensis* | B-1668 | negative | *Pseudomonas aeruginosa* | B-0040 | negative |
| *Campylobacter jejuni* | B-1229 | negative | *Serratia marcescens* | B-0014 | negative |
| *Candida albicans* | B-1266 | negative | *Sphingomonas zeae* | JM-791 | negative |
| *Citrobacter freundii* | B-0022 | negative | *Staphylococcus aureus* | B-0946 | negative |
| *Clostridium paraperfringens* | B-1435 | negative | *Staphylococcus epidermidis* | B-0026 | negative |
| *Clostridium sporogenes* | B-1450 | negative | *Stenotrophomonas maltophila* | B-0055 | negative |
| *Eikenella corrodens* | B-0614 | negative | *Streptococcus pneumoniae* | B-0847 | negative |
| *Escherichia coli* | B-1324 | negative | *Streptococcus pyogenes* | B-0846 | negative |
| *Francisella tularensis holarctica* | F-0049 | negative | *Vibrio cholerae* | B-1302 | negative |
| *Haemophilus influenzae* | B-0850 | negative | *Yersinia enterocolitica* | B-0099 | negative |
| *Klebsiella pneumoniae* | B-0008 | negative | *Yersinia pestis* | EV-76 | negative |
| *Legionella pneumophila* | B-1341 | negative | *Monkey Pox Virus* | MSF-6 | negative |
| *Listeria monocytogenes* | DSM-12464 | negative | *Vaccinia Virus* | VACV-0273/2004 | negative |
| | | | *Varicella Zoster Virus* | none | negative |

* results of n = 3 tests.

Supplementary Table S13: Linearity of the 16S rRNA SNP-PCR*.

| [Template]** | Assays day 1 | | | Assays day 2 | | | Assays day 3 | | | (Ø±SD)*** |
|---|---|---|---|---|---|---|---|---|---|---|
| **A.** | | | | | | | | | | |
| $10^9$ | 9.3 | 9.4 | 9.5 | 9.7 | 9.8 | 9.8 | 9.1 | 9.1 | 9.1 | 9.4±0.3 |
| $10^8$ | 12.6 | 12.6 | 12.5 | 13.0 | 13.1 | 13.0 | 12.3 | 12.3 | 12.3 | 12.6±0.3 |
| $10^7$ | 16.0 | 16.0 | 16.0 | 16.7 | 16.6 | 16.6 | 15.8 | 15.8 | 15.9 | 16.1±0.3 |
| $10^6$ | 19.2 | 19.1 | 19.0 | 19.5 | 19.6 | 19.6 | 18.8 | 18.8 | 18.8 | 19.2±0.3 |
| $10^5$ | 22.6 | 22.7 | 22.6 | 23.0 | 23.1 | 23.2 | 22.5 | 22.3 | 22.3 | 22.7±0.3 |
| $10^4$ | 25.9 | 25.8 | 26.0 | 26.2 | 26.3 | 26.3 | 25.6 | 25.6 | 25.6 | 25.9±0.3 |
| $10^3$ | 29.5 | 29.4 | 29.6 | 30.0 | 29.9 | 29.9 | 29.2 | 29.1 | 29.0 | 29.5±0.3 |
| $10^2$ | 32.3 | 32.9 | 32.3 | 32.9 | 33.0 | 32.8 | 32.4 | 32.2 | 32.0 | 32.5±0.3 |
| $10^1$ | 35.8 | 35.7 | 36.2 | 36.1 | 35.8 | 35.8 | 35.9 | 36.2 | 35.8 | 35.9±0.2 |
| $10^0$ | 36.9 | 37.3 | 38.8 | 39.2 | 38.9 | 38.6 | - | 37.2 | - | 38.1±1.0 |
| **B.** | | | | | | | | | | |
| $10^7$ | 17.1 | 17.0 | 17.1 | 16.7 | 16.8 | 16.8 | 16.8 | 16.8 | 16.8 | 16.9±0.2 |
| $10^6$ | 20.4 | 20.7 | 20.4 | 20.2 | 20.2 | 20.2 | 20.3 | 20.3 | 20.3 | 20.3±0.2 |
| $10^5$ | 24.0 | 24.0 | 24.1 | 23.8 | 23.7 | 23.7 | 23.8 | 23.7 | 23.9 | 23.9±0.2 |
| $10^3$ | 27.4 | 27.3 | 27.5 | 26.9 | 27.0 | 26.8 | 27.0 | 27.1 | 27.1 | 27.1±0.2 |
| $10^2$ | 30.8 | 29.6 | 30.7 | 30.5 | 30.5 | 30.6 | 30.5 | 30.7 | 30.4 | 30.5±0.3 |
| $10^1$ | 34.0 | 34.1 | 33.8 | 33.7 | 33.7 | 33.8 | 33.6 | 34.0 | 33.9 | 33.8±0.2 |
| $10^0$ | 38.3 | 36.7 | 37.1 | 36.1 | 36.9 | 36.7 | 37.3 | 37.1 | 36.6 | 37.0 ±0.6 |

* Replicates (n=3) of each PCR reaction with target DNA were run on three consecutive days; ** templates at indicated concentration per reaction were cloned fragment (A) or *B. anthracis* Ames DNA (B); *** averages (Ø) ± standard deviations (SD) of n = 3 x 3 tests.

Supplementary Table S14: Probit-analysis of the 16S rRNA SNP-PCR.

| Copies/reaction | # of tests | # of positive tests |
|---|---|---|
| 10 | 12 | 12 |
| 8 | 12 | 12 |
| 6 | 12 | 12 |
| 4 | 12 | 12 |
| 3 | 12 | 12 |
| 2 | 12 | 7 |
| 1 | 12 | 4 |

Supplementary Table S15: Linearity of the 16S rRNA SNP RT-PCR*.

| [Template]** | Triplicates | | | (Ø±SD)*** |
|---|---|---|---|---|
| $10^8$ | 10.2 | 9.8 | 10.4 | 10.1±0.3 |
| $10^7$ | 13.7 | 13.7 | 13.8 | 13.7±0.1 |
| $10^6$ | 16.7 | 16.7 | 16.8 | 16.7±0.1 |
| $10^5$ | 20.0 | 19.5 | 21.2 | 20.2±0.9 |
| $10^4$ | 24.5 | 24.3 | 23.8 | 24.2±0.3 |
| $10^3$ | 26.6 | 26.9 | 26.6 | 26.7±0.2 |
| $10^2$ | 29.8 | 31.2 | 30.9 | 30.6±0.7 |
| $10^1$ | 34.7 | 35.0 | 34.9 | 34.8±0.2 |
| $10^0$ | 40.3 | 37.0 | 40.0 | 39.1±1.8 |

* Replicates (n=3) of each PCR reaction with target DNA were run; ** templates at indicated concentration per reaction were RNAs (including genomic) DNA of *B. anthracis* Sterne; *** averages (Ø) ± standard deviations (SD) of n = 3 tests.

Supplementary Table S16: Probit-analysis of the 16S rRNA SNP RT-PCR.

| Copies/reaction | # of tests | # of positive tests |
|---|---|---|
| 15 | 12 | 12 |
| 12 | 12 | 12 |
| 9 | 12 | 12 |
| 7.5 | 12 | 12 |
| 6 | 12 | 10 |
| 4.5 | 12 | 10 |
| 3 | 12 | 8 |
| 1.5 | 12 | 8 |
| 0 | 12 | 0 |

### *6.2. Supplementary Figures*

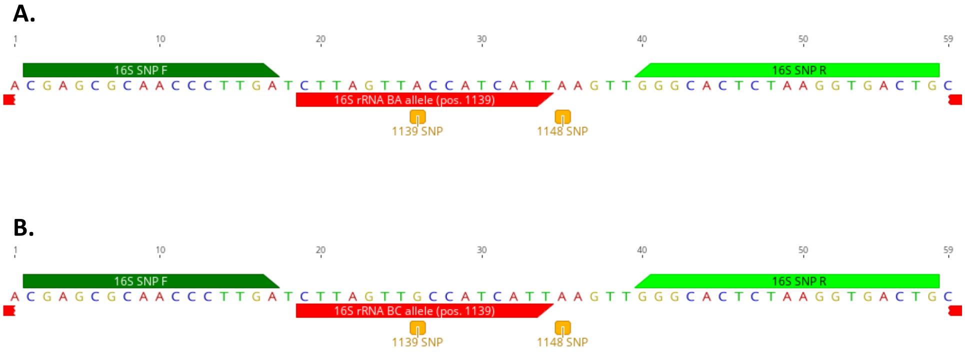


Supplementary Figure S1: Partial sequence of different alleles of the 16S rRNA genes of *B. anthracis* with specific SNP ("A" at position 1139 according to (3), position 1110 in *B. anthracis* Ames Ancestor NC_007530)* and primer and probe positions. (A) *B. anthracis*-specific 16S-BA-allele. (B) alternative 16S rRNA gene allele (16S-BC-allele) in *B. anthracis* common for the *B. cereus*-group with alternative SNP state ("G" at position 1110 in *B. anthracis* strain Ames Ancestor, NC_007530). *a second SNP at position 1119 present in some of the 16S rRNA gene alleles was found not to be specific for *B. anthracis* (3).

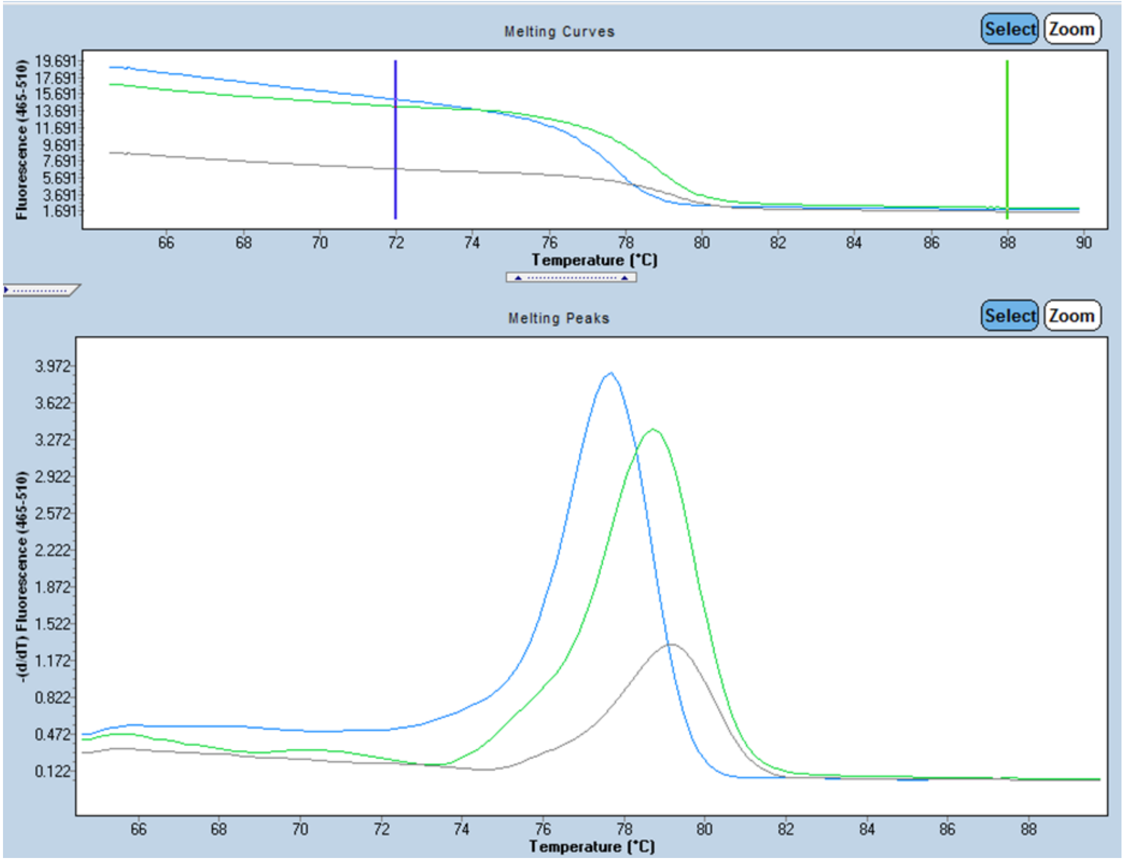


Supplementary Figure S2: Melting point analysis of PCR products. Shown are melting point analyses as fluorescence change (upper panel) or as its first derivative (lower panel) over a temperature gradient. Amplificates were 16S-BA-allele (100 template copies, blue line), 16S-BC-allele (100 template copies, green line) or water-only control (grey line).

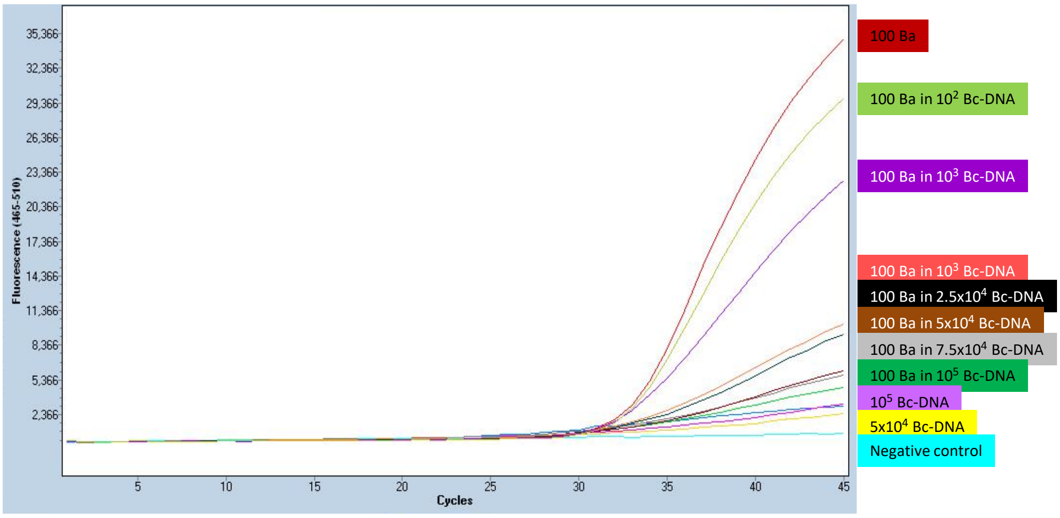


Supplementary Figure S3: Competitive inhibition the 16S rRNA SNP-PCR assay by the alternative 16S-BC-allele. A constant 100 template copies of the 16S-BA-allele per reaction were titrated against increasing copy numbers of the alternative 16S–BC-allele and fluorescence recorded (shown are representative curves; see Supplementary Table S9 for numerical data as replicates).

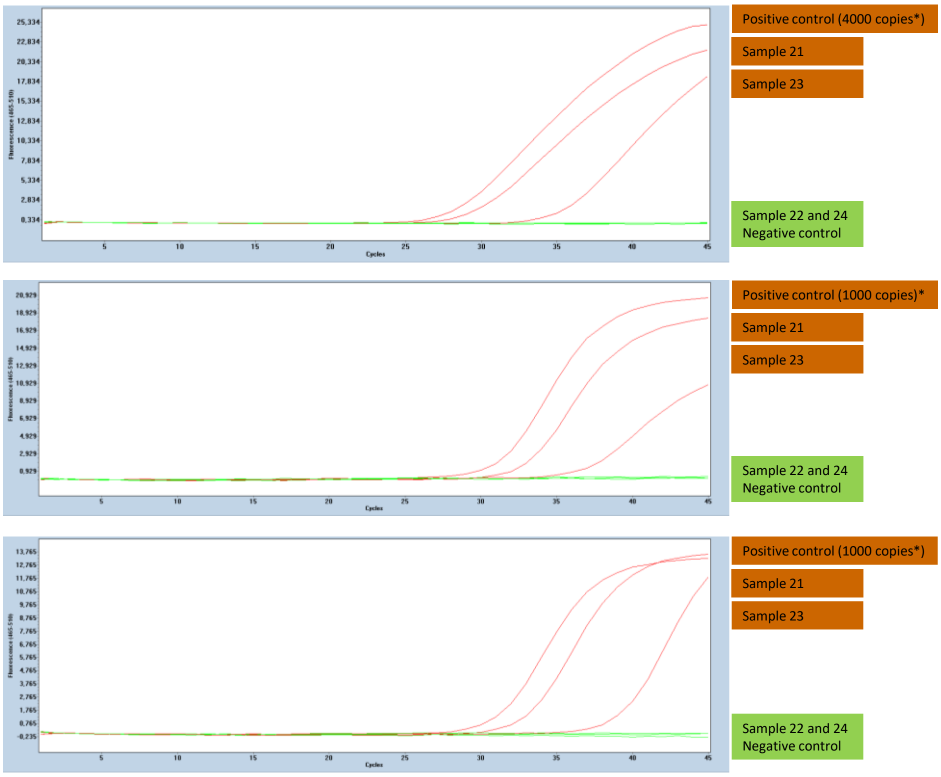


Supplementary Figure S4: Challenge of the new 16S rRNA-based multi-copy assay with samples from a ring trial. Samples were subjected to real time PCR using the new 16S rRNA SNP assay (upper panel), published *dhp61* gene assay (4) (middle panel) or published *PL3* gene assay (5) (lower panel). Representative amplification curves (from n=3 with similar results) are shown. *Positive controls were 1000 genomes of *B. anthracis* Ames (harboring 4000 copies of the 16S-BA-allele but only 1000 copies of *dhp61* or *PL3*, respectively.

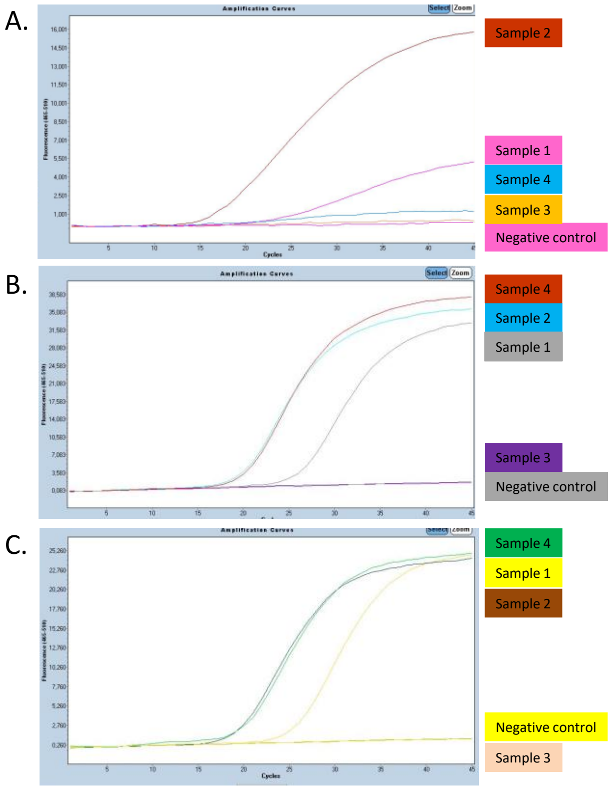


Supplementary Figure S5: Challenge of the new 16S rRNA-based multi-copy assay with total DNA from spiked soil. Samples were subjected to real time PCR using the new 16S rRNA SNP assay (upper panel), published *dhp61* gene assay (4) (middle panel) or published *PL3* gene assay (5) (lower panel). Representative amplification curves (from n=3 with similar results) are shown.
